# Exhaled aerosol increases with COVID-19 infection, and risk factors of disease symptom severity

**DOI:** 10.1101/2020.09.30.20199828

**Authors:** David A. Edwards, Dennis Ausiello, Robert Langer, Jonathan Salzman, Tom Devlin, Brandon J. Beddingfield, Alyssa C. Fears, Lara A. Doyle-Meyers, Rachel K. Redmann, Stephanie Z. Killeen, Nicholas J. Maness, Chad J. Roy

## Abstract

Coronavirus disease-19 (COVID-19) transmits by droplets generated from surfaces of airway mucus during processes of respiration within hosts infected by severe acute respiratory syndrome-coronavirus-2 (SARS-CoV-2) virus. We studied respiratory droplet generation and exhalation in human and nonhuman primate subjects with and without COVID-19 infection to explore whether SARS-CoV-2 infection, and other changes in physiological state, translates into observable evolution of numbers and sizes of exhaled respiratory droplets in healthy and diseased subjects. In our observational cohort study of the exhaled breath particles of 74 healthy human subjects, and in our experimental infection study of eight nonhuman primates infected by aerosol with SARS-CoV-2, we found that exhaled aerosol particles increase one to three orders of magnitude with aging, high BMI, and COVID-19 infection. These variances appear to be related to changes in airway mucus surface composition and the propensity for mucus surfaces to breakup into small droplets during acts of breathing. We also observed that 20% of those participating in our human study accounted for 80% of the overall exhaled bioaerosol, reflecting a bioaerosol distribution analogous to a classical 20:80 super spreader distribution.

## INTRODUCTION

Severe acute respiratory syndrome-coronavirus-2 (SARS-CoV-2) transmits through the air by a combination of the large droplets exhaled when people cough or sneeze, and by the very small droplets people generate in their airways when they naturally breathe (1-4). How exhaled respiratory droplets vary between individuals, evolve over time within individuals, and change with the onset and progression of COVID-19 infection is poorly understood, yet critical to clarifying the nature of COVID-19 transmission — and other highly-communicable airborne respiratory diseases, such as influenza and tuberculosis.

Generation of respiratory droplets in exhaled breath can occur by the force of the fast air flows that occur in the upper airways when we breathe, talk, cough and sneeze. At peak inspiratory flows during normal breathing air speeds in the trachea and main bronchi can reach turbulent velocities (5). The rush of air over the thin (5 to 10 μm) mucus layer lining the airways can break up the mucus surface into small droplets in the way strong winds produce breakup and spray on the surface of the ocean (6). The nature and extent of this droplet breakup is dependent on the surface properties of the mucus itself (6, 7). Among properties most influencing droplet generation and droplet size are surface viscoelasticity (which resists the stretching of mucus surface on breakup) and surface tension (which lowers the energy expended in small droplet creation) (8, 9). In airway lining mucus, both properties vary with lung surfactant type and concentration, as well as with composition and structure of mucus in close proximity to air surfaces (6). Surfactant and mucin compositional and structural changes, driven in part by physiological alterations of the human condition — including diet (10), aging (11), and COVID-19 infection itself (12) — may therefore be anticipated to alter droplet generation and droplet size (7) during acts of breathing.

To ascertain whether COVID-19 infection and other phenotypical differences associated with severity of infection risk might alter airborne droplet generation from airway lining fluid during acts of breathing, we conducted two studies in human and nonhuman primates. In our first study, we evaluated the exhaled breath of 74 human subjects to determine exhaled breath particle variations in the human population. In our second study, we measured the exhaled breath from two species of nonhuman primates following experimental infection via inhalation of SARS-CoV-2. We then assessed exhaled breath particle evolution over the time-course of exhaled aerosol particles as a function of nasal viral titer. We report on these studies here.

## RESULTS & DISCUSSION

We evaluated the exhaled aerosol of 74 human volunteers on two consecutive days in an observational cohort study of essential workers at No Evil Foods in Asheville, North Carolina. The results from these measurements (Figure 1A), with each volunteer represented as two subjects (days one and two).

**Figure 1.**
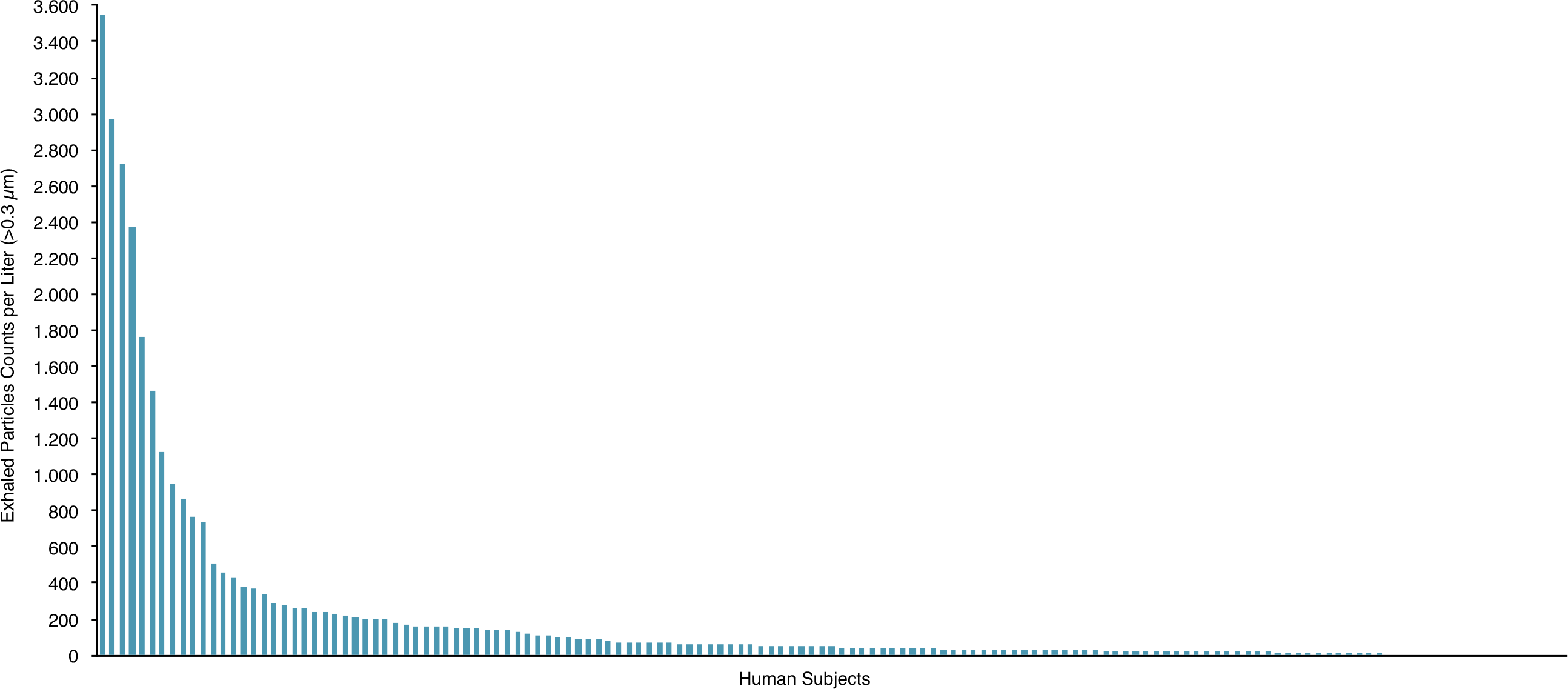
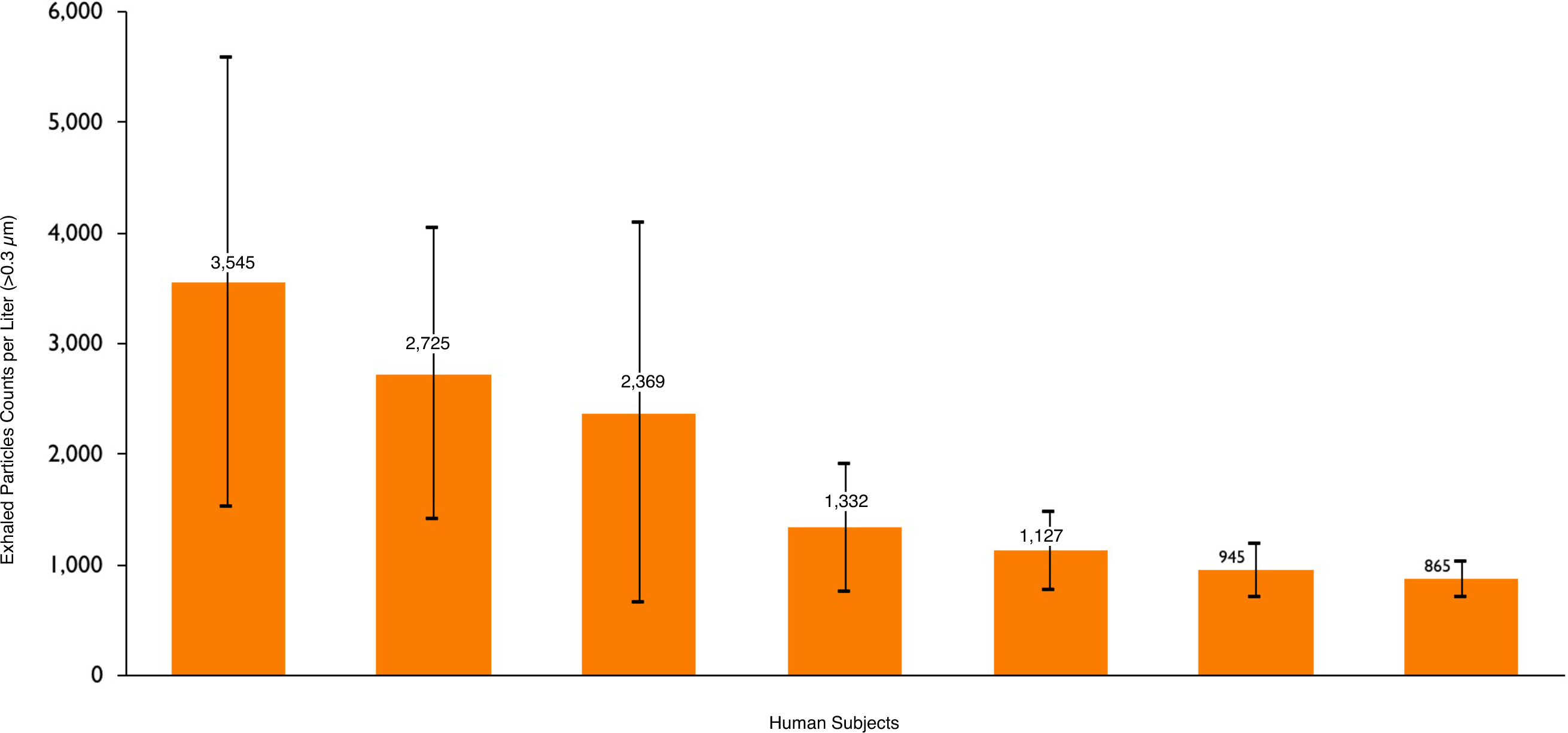
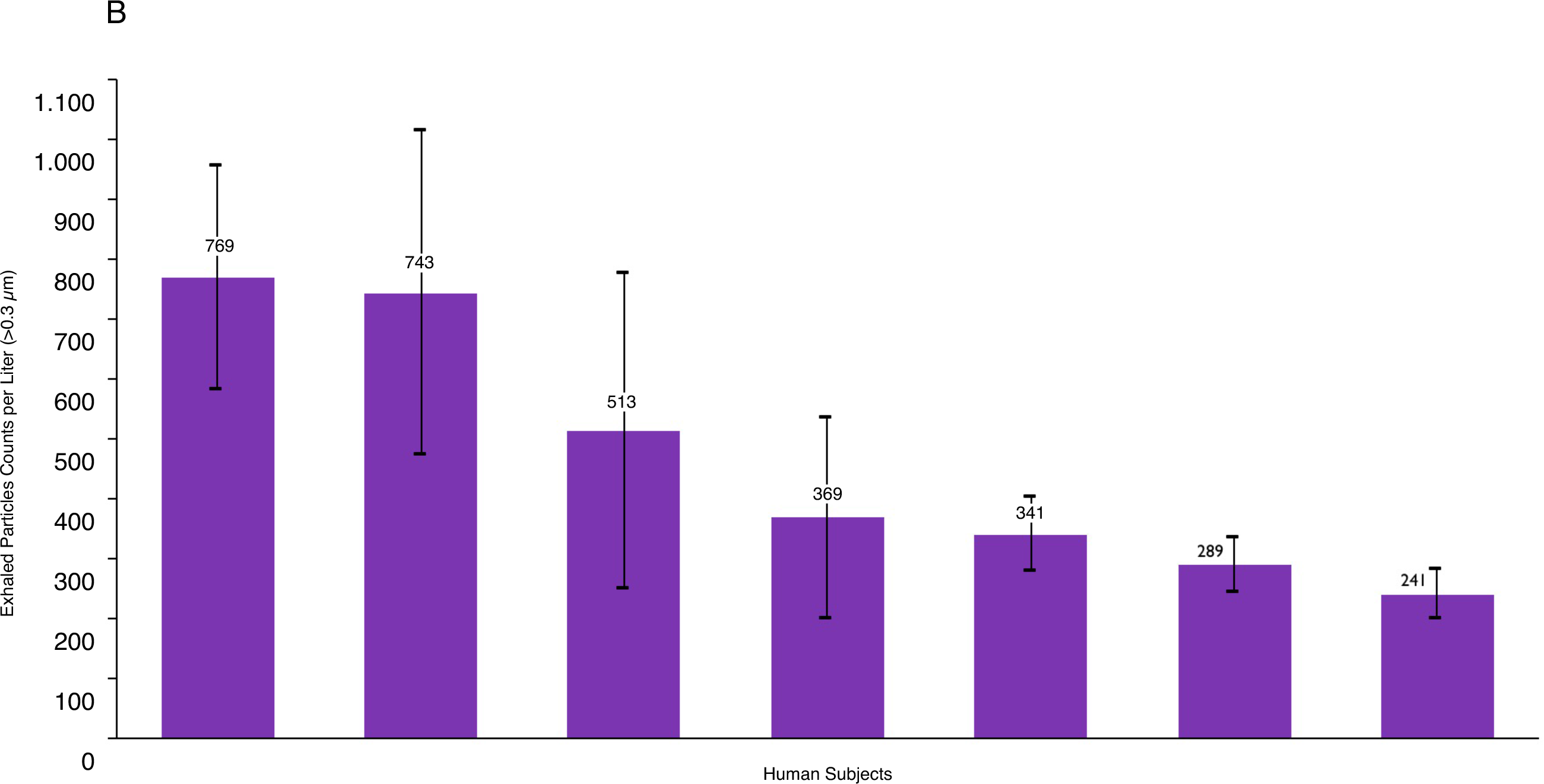
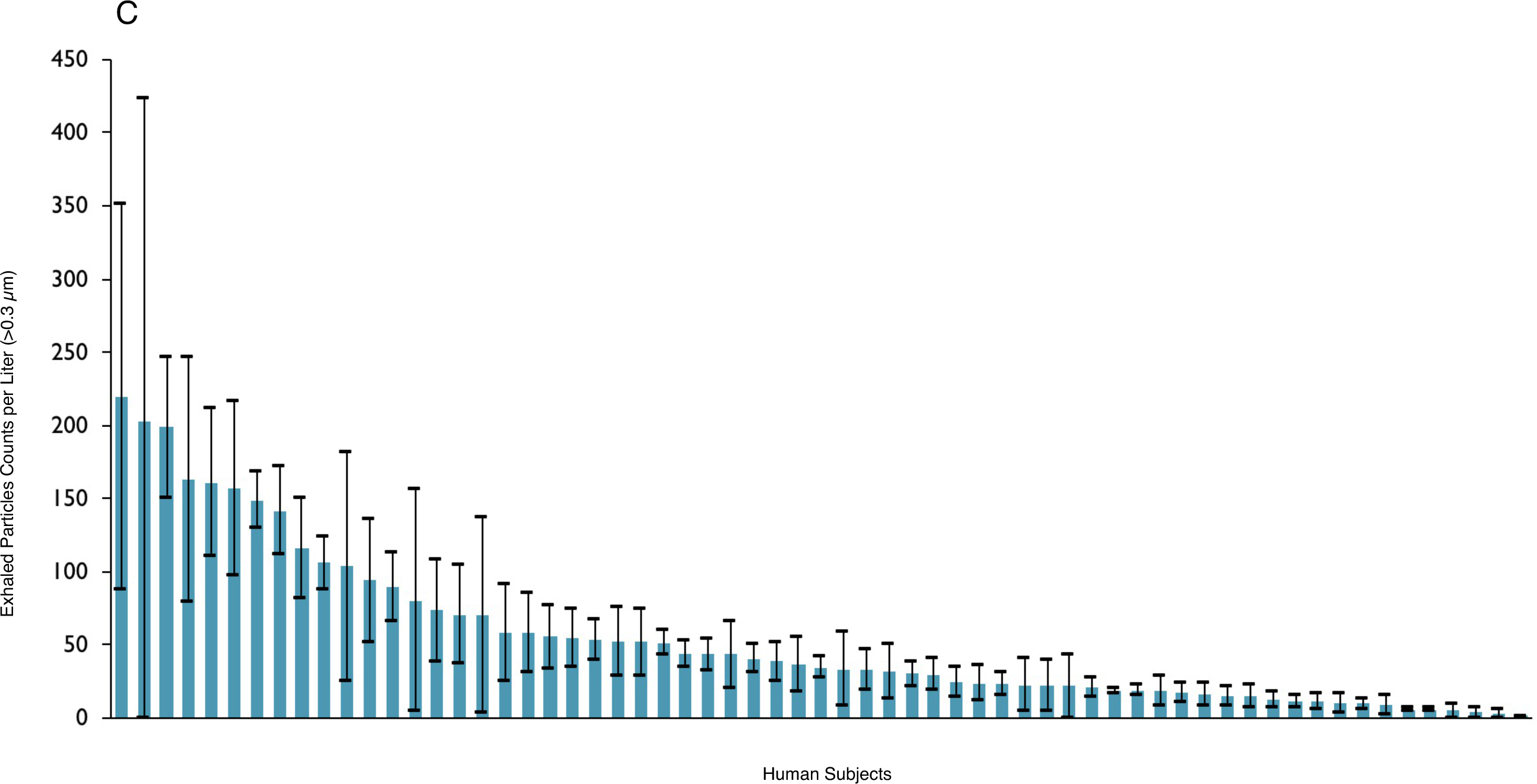
Exhaled breath particles of 74 essential workers at No Evil Foods. **A**. All participants on days 1 & 2; **B** “Super Spreader” participants day one (1st decile); **C** “Super Spreader” participants day one (2nd decile); **D** “Low Spreader” participants day one. Data represent particle counts per liter of exhaled air (particle diameter larger than 300 nm) for each of the 74 individuals.

We categorized subjects by those exhaling greater or less than 240 particles per liter of air. We chose this demarcation since the individuals above this threshold aerosol number exhaled 80% of the total particle production on day one while being only 20% of the total members of the group — analogous to the conventional definition of super spreading of airborne infectious disease (13). Within this high producing group, we noted that 80% of the super spreader production was generated by half of the group, i.e. seven individuals. We qualified as “low spreaders” those 60 individuals who exhaled below 240 particles per liter. The individual data for each category are shown with standard deviations (day one only; Figure 1B-D). Exhaled aerosol particle numbers varied appreciably over the course of breathing per individual and in proportion to the overall exhaled aerosol particle number.

We evaluated relationships between exhaled aerosol particle number and sex, age, and body mass index (BMI). No correlation was found with sex, while significant correlation between exhaled aerosol and age and BMI was observed. All volunteers <26 years of age (for whom we collected age and BMI information) exhaled fewer particles (low spreaders; Figure 1B) than those above 26 years of age (p<0.00010), while all volunteers (of any age) with BMI less than 22 exhaled fewer particles (low spreaders) than those with BMI above 22 (Figure 1C) (p<0.0069). For older subjects and those of higher BMI the likelihood of exhaling higher numbers of particles on average tended to increase with advanced age and BMI (Figures 1B, 1C). The coupled effects of age and BMI (Figure 1D) are in terms of BMI-years (Age-BMI multiple). All participants with BMI-years <950 have lower numbers of exhaled particles than those a >950 BMI-years (p<0.0362).

To explore the dependence of exhaled aerosol on COVID-19 infection we studied the course of infection in a nonhuman primate model. Two species of nonhuman primate (*Macaca mulatta*, rhesus macaque; and *Chlorocebus aethiops*; African green monkey) were experimentally infected with SARS-CoV-2 by small particle aerosol (≈2 µm) and closely monitored thereafter. Results of mucosal sampling (nasal swabs) showed productive infection in both species, with viral RNA detected as early as +1 day post-infection; viral titers reaching a crescendo in most animals by day +7 post infection, and clear declination by day +14 post infection, and to undetectable concentrations by study terminus (+28 days post infection).

The results of the exhaled breath particle production followed a remarkably similar temporal pattern as SARS-CoV-2 viral replication measured in the nasal swabs (Figure 3). Total exhaled breath particle production began to increase starting at +3 day post infection and continued to rise by day 7, and decreased to essentially baseline levels by day 14 in both species in all animals across both species.

**Figure 2.**
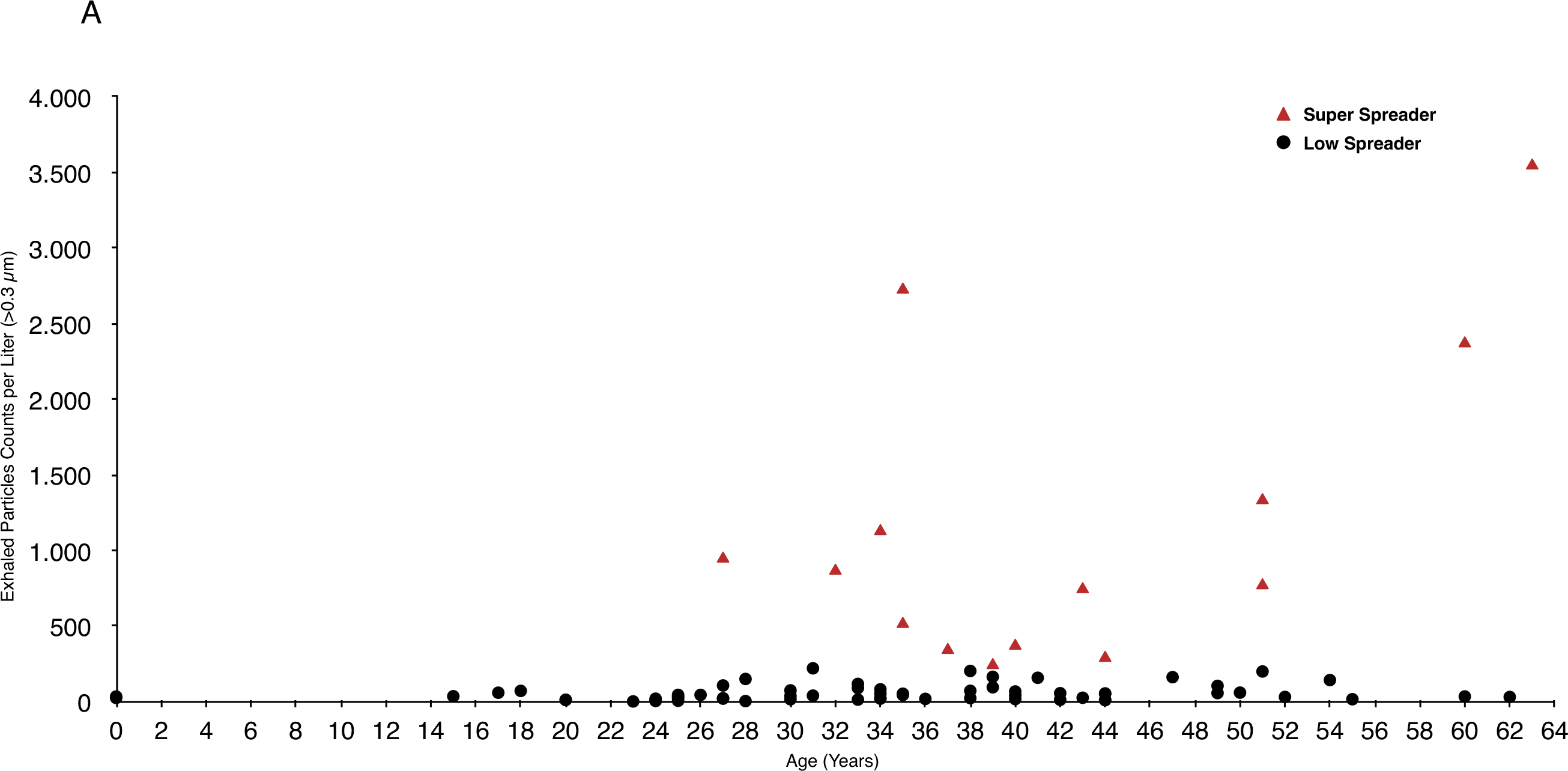
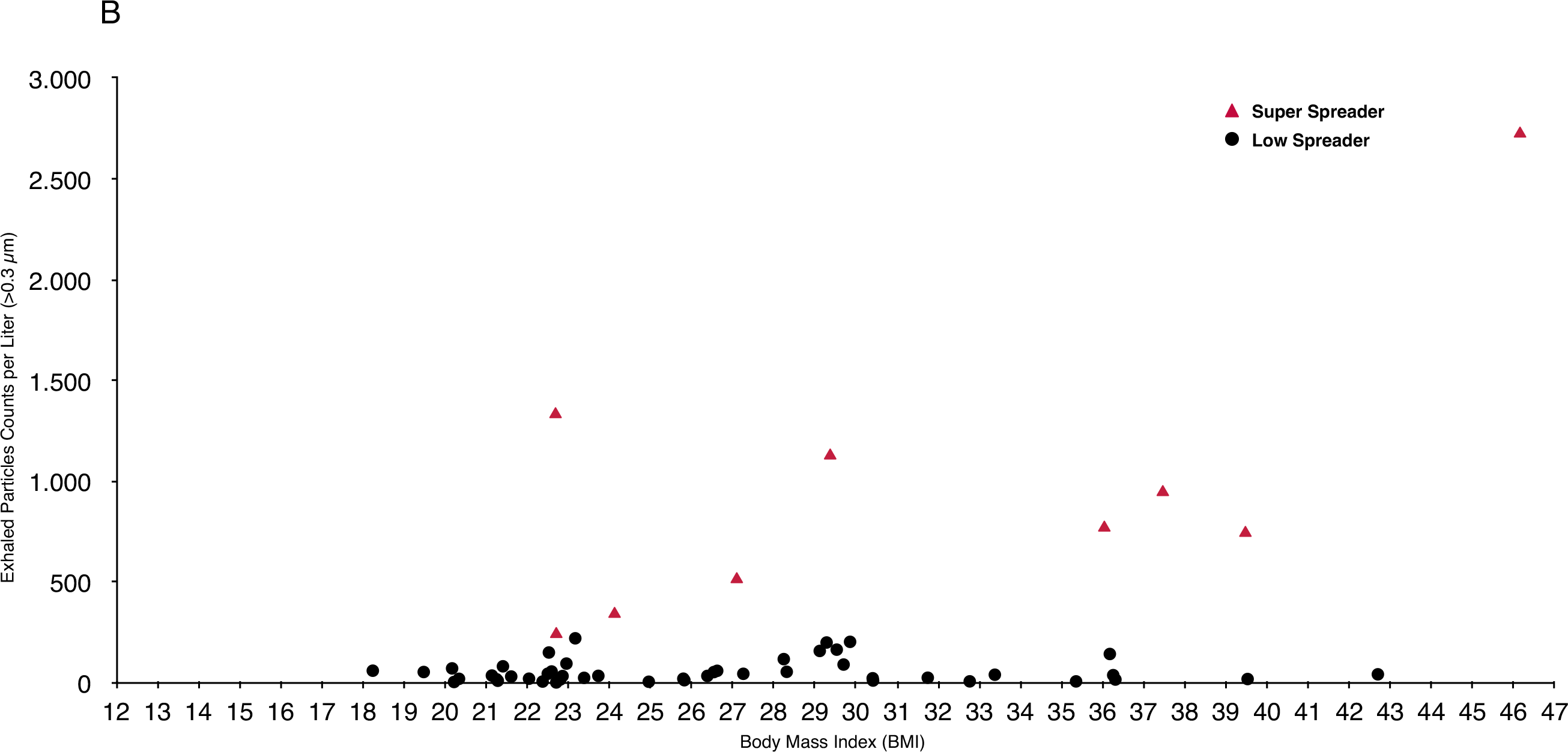
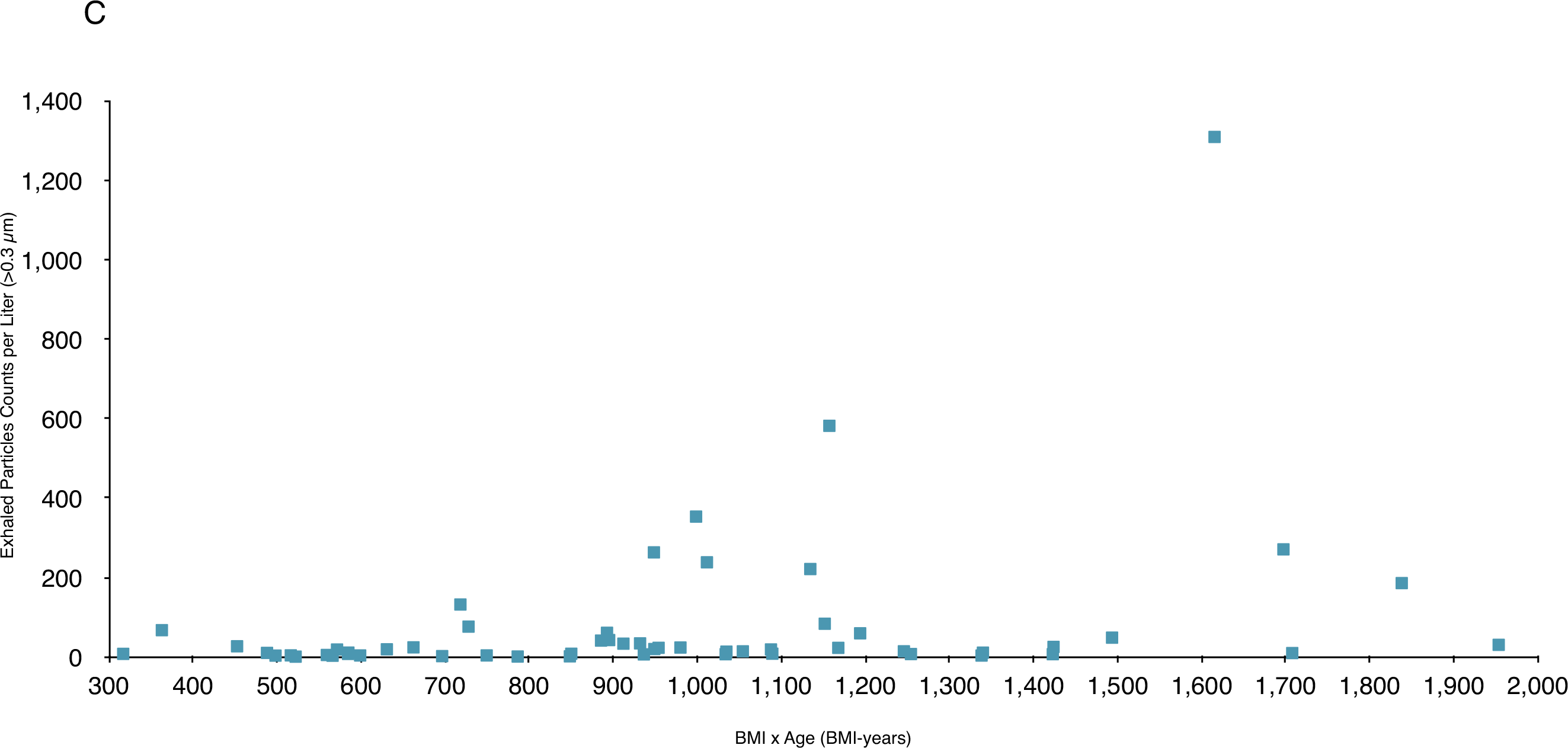
Exhaled breath particles of volunteers reporting age and BMI: **A v**ersus age; **B v**ersus BMI; **C** versus BMI-years. Data represent particle counts per liter of exhaled air (particle diameter larger than 300 nm) for each of the 74 individuals on day one. Error bars represent standard deviation sample calculations based on three to 12 exhaled aerosol count measurements with each measurement an average of counts over a five second time interval.

**Figure 3.**
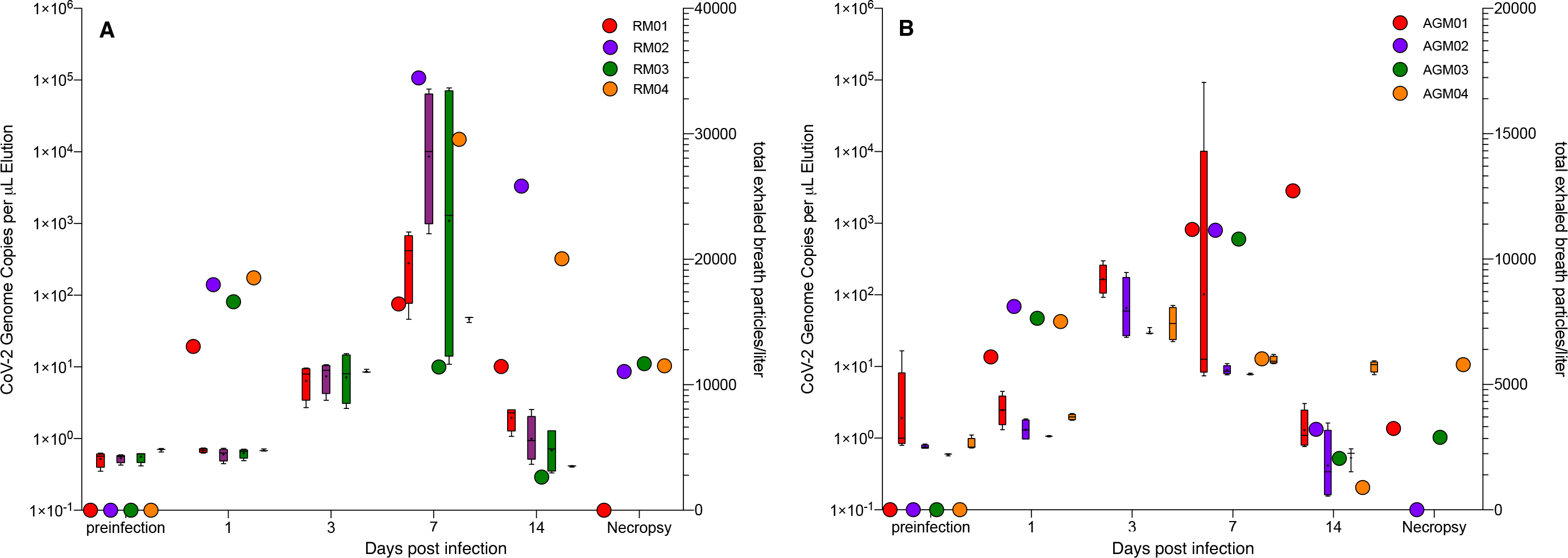
Exhaled breath particles and corresponding genomic SARS-CoV-2 viral RNA in experimentally-infected Rhesus macaques (RM; panel **A**) and African green monkeys (AGM; panel **B**). Both groups segregated by species (n=4; N=8). The corresponding color-matched box-and-whisker plots of total exhaled breath particles represent iterative five (5) one-minute sampling events to genomic viral RNA (color-matched circles) for each animal at each respective timepoint. Mean calculated correlation between timepoint-matched exhaled breath particle production and genomic viral RNA showed statistically significant correlations in 75% of the RM (RM01, r2=0.93, *p*<0.03; RM02, r2=0.99, *p*<0.004; RM04, r2=0.98, *p*<0.0008) and 50% of the AGM (AGM02, r2=0.91, *p*<0.04; AGM03, r2=0.97, *p*<0.01).

Although increase of total particles was observed in both species, aerosol particle production increase relative to pre-infection totals was more profound in the Rhesus macaques (Figure 3A) than in the African green monkeys (Figure 3B). There was a statistically significant correlation between the production of exhaled breath particles and corresponding nasal viral genomic RNA in 75% of the Rhesus macaques (RM01, r^2^=0.93, *p*<0.03; RM02, r^2^=0.99, p<0.004; RM04, r^2^=0.98, *p*<0.0008) and 50% of the African green monkeys (AGM02, r^2^=0.91, *p*<0.04; AGM03, r^2^=0.97, *p*<0.01).

The particle distributions of the exhaled breath particles, averaged among species-segregated cohort, changed as COVID-19 disease progressed (Figure 4A, B). The shift in particle size is typified by a clear increase of particles categorized as <1 µm (collective of the 3 size bins of 0.3, 0.5, and 1.0 µm) initiating on day +3 post infection and continuing to trend to a smaller particle size by day seven, with a slight rebound of particle size distribution to baseline by day 14. Collectively, the total number and the relative size distribution of exhaled breath particles produced during experimentally-induced COVID-19 is correlated with the viral kinetics of SARS-CoV-2 infection in both the Rhesus macaque and African green monkey nonhuman primate species.

**Figure 4.**
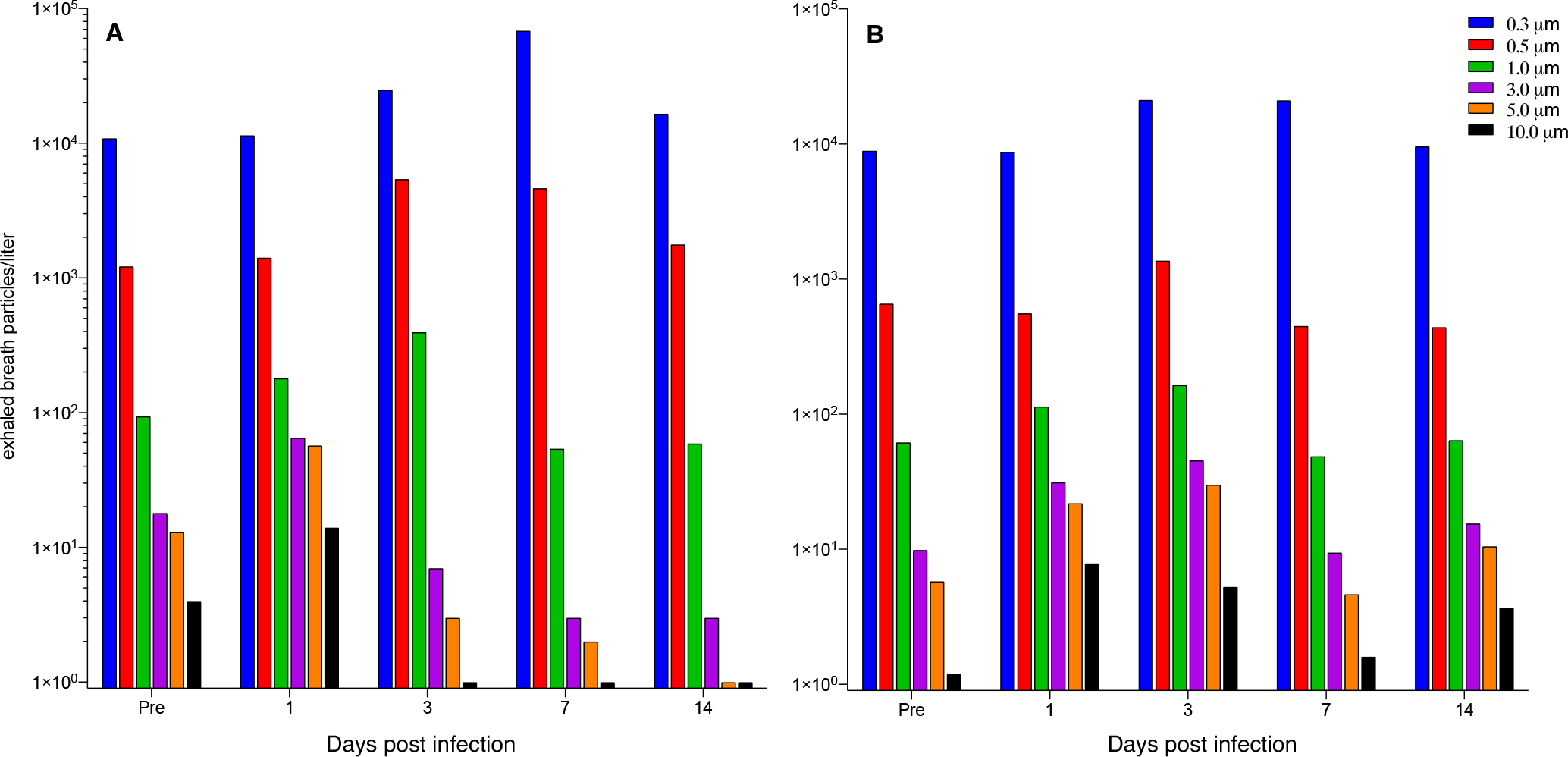
Exhaled breath particles and corresponding particle size distributions in experimentally-infected Rhesus macaques (RM; panel A) and African green monkeys (AGM; panel B). **Figure 6**. Production of exhaled breath particles and corresponding particle size distributions in experimentally-infected Rhesus macaques (RM; panel **A**) and African green monkeys (AGM; panel **B**). Distribution bars represent mean of total particle count at that particular size cutoff (legend) for each species group (n=4).

The combined results of our two studies reveal that numbers of exhaled aerosol particles vary with COVID-19 infection and risk factors (advanced age, high BMI) for severity of COVID-19 symptoms. That exhaled breath particles vary substantially between individuals in an uninfected human population (Figure 1A), and adhere to classical super spreading 20:80 rules (13), further suggests that the nature of COVID-19 transmission, and super spreading (14, 15), is not only a matter of air currents and proximity of infected and naive hosts, but also of human phenotype.

While the results of our studies suggest that some people are especially at risk of producing large numbers of exhaled aerosols, they also indicate that most people can be at risk for virus penetration deep into the lungs, and exhalation of virus into the environment, following upper airway deposition of SARS-CoV-2. It seems nevertheless probable that those with higher propensity to exhale larger numbers of particles, like the elderly, once infected may be most likely to transmit the disease, underlining the special risk of living circumstances that bring high-risk populations into close proximity for extended periods of time, such as nursing homes.

We found that the proportion of small respiratory droplets (the majority of particles exhaled in all subjects) increased at the peak of COVID-19 infection in nonhuman primates (Figure 4). This diminution in exhaled aerosol particle size indicates that at peak infection there may be an elevated risk of the airborne transmission of SARS-CoV-2 by way of the very small droplets that transmit through conventional masks and traverse distances far exceeding the conventional social distance of two meters.

The scientific response to the COVID-19 pandemic has largely focused on the development of curative drugs and preventive vaccines. In the wait for a cure or an effective widely adopted vaccine, it may be advisable for the scientific community to additionally focus on management of COVID-19 — in part through the diminution of exhaled breath particles. Beyond more effective masks and more sophisticated social distancing rules, approaches to stabilize airway lining mucus and retain mucus clearance function might be particularly useful.

Exhaled aerosol numbers appear to be not only an indicator of disease progression, but a marker of disease risk in non-infected individuals. Monitoring (as a diagnostic) might also be an important strategy to consider in the control of transmission and infection of COVID-19 and other respiratory infectious diseases, including influenza.

## METHODS

### Trial Design and Participants

We conducted an observational cohort human volunteer study designed to evaluate exhaled aerosol particle size and number during normal breathing in non-infected humans. In the conducting of the study and the reporting of our results we followed Strengthening the Reporting of Observational Studies in Epidemiology (STROBE) statement reporting guidelines. Eligible participants were healthy adults 19 to 66 years of age, all essential workers at No Evil Foods in Asheville North Carolina. Participants were not screened for SARS CoV-2 infection by serology or polymerase chain reaction (PCR) before enrollment. The trial was conducted on the premises of No Evil Foods. The protocol is available in the Supplementary Appendix. An independent review board (Ethical and Independent Review Services) determined formal Institutional Review Board (IRB) review to be unnecessary when considering the observational nature of the study the corresponding minimal impact on human subject research.

### Study Procedures

Participants spent up to 30 minutes per session during their quarantined home routine or away from work or school to have their exhaled aerosol particles measured. Exhaled particles were measured by a particle detector (Climet 450-t) designed to count airborne particles in the size range of 0.3 to 5 μm. The particle detector was connected to standard nebulizer tubing and mouthpiece that filters incoming air through a HEPA filter. Each standard nebulizer tubing and mouthpiece were removed from sealed packaging before each subject prior to the subject’s first exhaled particle detection. On subsequent counting maneuvers the same mouthpiece, tubing and HEPA filter were replaced into the particle counter system by the participant to insure effective hygiene. Subjects performed normal tidal breathing through a mouthpiece while plugging their noses over one to two minutes — beginning with two deep breaths to empty their lungs of environmental particles. Over this time frame particle counts per liter diminished to a lower baseline number reflecting particles emitted from breakup of airway lining fluid surfaces in the subject’s airways. Once the lower plateau of particle counts was reached subjects continued to breathe normally. Three to eight particle counts (average values of particle counts assessed over six seconds) were then averaged to determine the mean exhaled particle count and standard deviation. Participants sat opposite to the study administrator with a plexiglass barrier separating the participant and the administrator.

### Nonhuman Primate Experimental SARS-CoV-2 Infection

#### Animals and procedures

A total of eight male (>7 years of age), purpose-bred Rhesus macaques and wild caught African green monkeys were used in our nonhuman primate study. Animals were exposed to SARS-CoV-2 (BEI, USA-WA1/2020, NR-52281) by small particle aerosols previously characterized in our laboratory inhalation system (16). Animals received an inhaled dose of ∼2.5×10^3^ TCID50 (Tissue Culture Infectious Dose 50; supplemental data). Animals were observed for 28 days post infection including twice daily monitoring by veterinary staff. Mucosal and other biosamples were collected at seven days before infection, 1, 3, 7, 14, and at necropsy (day 28) after infection. During biosampling events and physical examination while anesthetized and in dorsal recumbency, and experiencing normal respiration, animals were individually sampled for exhaled breath aerosols. This sampling was performed using a modified pediatric face mask fitted with a HEPA-filtered inspiration port, and corresponding sampler for exhalation. A particle counter (Thermo Systems Inc. (TSI) AeroTrak handheld particle counter Model 9306-V2, St. Paul, MN) was used to sample exhaled breath particles for five one-minute intervals at every sampling time point. Exhaled breath particle data were collected in a cumulative fashion.

#### Quantification of Swab Viral RNA

Nasal swabs were collected in 200 µL of DNA/RNA Shield and extracted for Viral RNA (vRNA) using the Quick-RNA Viral kit. Samples were then quantified using RT-qPCR (Supplemental Methods).

#### Ethics

The Institutional Animal Care and Use Committee of Tulane University reviewed and approved all the procedures for this experiment. The Tulane National Primate Research Center is fully accredited by the Association for Assessment and Accreditation of Laboratory Animal Care. All animals are cared for in accordance with the NIH guide to Laboratory Animal Care. The Tulane Institutional Biosafety Committee approved the procedures for sample handling, inactivation, and removal from biosafety level 3 (BSL-3) containment.

## Data Availability

Data and Materials Availability. All data from this study are presented in the article and Supplementary Materials.

## ACKNOWLEDGMENTS

The authors would like to thank the leadership and all employees of No Evil Foods for their participation in this study. The authors would also like to thank Stephanie and her family for their willingness to participate in this study during their home quarantine.

## Author contributions

DE, DA, RL conceived and designed the human research, and CJR conceived and designed the studies involving nonhuman primates. DE, JS and TD executed the human trials. BB, AF, LD, RR, SK and NM performed the nonhuman primate studies. DA and CJR wrote and edited the manuscript.

## Data and Materials Availability

All data from this study are presented in the article and Supplementary Materials.

