## Supplemental Materials for "Exhaled aerosol increases with COVID-19 infection, and risk factors of disease symptom severity"

**Document Number:** 01-0001
Version Number: 01.06

#### **Process Name:** Aerosol Assessment North Carolina Study

**Project Management Work Stream:** Research and Development

#### Contents

About This Document ix
1. Introduction
2. Scope
2.1. This process document contains information on the following:
 2.1.1.Suggested protocol for the measurement of exhaled and emitted aerosolized particles, by size of particles
 2.1.2.This version of the study focuses on the variations of particle aerosolization in participants over the course of multiple, closely timed measurements (e.g., daily over the course of a week, with multiple measurements taken each day).
 2.1.3.Additionally, this study design varies is other ways:
 2.1.4.Suggested directions for participants and administrators to effectively and consistently administer this process and inform participants of the test's mechanics to aid their understanding and successful participation of study requirements
 2.1.5.Required inventory list of equipment, supporting inventory, and study administration materials for successful deployment (see Appendix A)
3. References
3.1. Trial Flow Diagram: Trial Flow Layout Setup (See insert visuals in scope design listed below)
4. Terms and definitions
4.1. Nebulizer Kits
4.2. HEPA Filters and HEPA Filter Assembly
4.3. T-Connector Valve Caps (Used for HEPA Filter Testing / Baseline)
4.4. Particle Counter Receipt Paper
5. Roles and responsibilities
5.1. Study Project Manager
5.2. Study Site Point of Contact (POC)
5.3. Registration Administrator
5.4. Particle Test Administrator –
5.5. Runner to Coordinate with Participants 4
5.6. Trial participants
6. Process
6.1. Pre-Work / Things Done in Advance
 6.1.1.Gather/Acquire all materials required as per materials list (see full inventory list in Appendix A)
 6.1.2.Organize Dry-Run Before Trial

6.2. Visit Study Location Factory

6.2.1. Room Setup for Trial

6.3. Overview of Particle Exhalation Baseline

6.3.1. Participants will proceed through several measurements, taking place over several days. Please defer to the study Principle Investigator (i.e., PI) to determine the final protocol for the specific study administration.

6.3.2. In this study, participants within the same household will be tested at the same sequence.

6.4. Phase A Overview

6.5. Phase A: Registration

6.5.1. Participant comes into the test administration room. The Registration Administrator (Admin) welcomes them: Thank you for making time to participate in this study. Can I please have your First and Last Names?

6.5.2. Admin should also ask the participant if they have tested positive for COVID-19. If so, the administrator should make a note, and ask when the participant was tested, and when their symptoms were first observed.

6.5.3. Admin says Have you had a chance to review and sign our informed consent waiver?

6.5.4. Overview of process: The test process will take about 20 minutes. You will be asked to breathe into the particle counter machine using your own new filter kit assembly. We want to measure the number and size of particles our participants give off when you breath

6.5.5. Participant is handed their Ziploc bag with the mouthpiece inside. Admin says: At the next station, our Administrations will guide you through how to use the particle counter. To reduce the risk of any transmission of illness, only you will touch the component that will go in your mouth. This bag was packaged by our team, on clean surfaces, wearing gloves, to make sure it is clean for you.

6.5.6. The particle count test will run for about 5 minutes.

6.5.7. When you complete the test, our team will direct you back to me at the registration desk. At that time, I will collect and sanitize the bag with your Nebulizer Assembly (including your mouthpiece) to be kept by the desk

6.5.8. Do you have any questions before we begin?

6.5.9. Admin will hand the participant three items

6.5.10. Admin will say: Please proceed to the next station (indicating where it is). When you arrive, please give your envelope to the administrator at the station. It will have been sanitized before you arrive.

6.5.11. Admin can then proceed to address the following participant in line.

6.6. Phase A: Particle Exhalation

6.6.1. Before the participant arrives, the admin will come around from the particle counter and the separator screen. They will sanitize with wipes the seat, screen, and tabletop.

6.6.2. When the participant arrives, the admin will direct them to kindly sit down in the chair in front of the machine. Admin: Thank you for participating! At this station, we will measure the number and size of the particles you produce while breathing. At the registration table, you were given a bag and an envelope. Can I please have the envelope?

6.6.3. To begin, I will activate the machine so you get a sense for how it sounds and work. Right now, you will see the red cap is sealing where your mouthpiece and mouth will go. This will help us make sure the machine is properly working—and give us a baseline against which to measure your particle counts and sizes.

6.6.4. During this test, if the >0.3 µm count (the lowest of the four numbers) on the particle counter machine screen is > 40 for more than ten seconds, AND/OR if the>0.3 µm count is variable (i.e., the count jumps), let the machine run for 20 seconds (~3-4 6-second intervals). If the count continues to exceed the number above, or is variable, the admin should take the following steps to remediate:

6.6.5. If the numbers do not exceed thresholds, the Admin should turn off the counter (but hitting STOP on the bottom left hand yellow button) for machine. The test can begin. The Admin will then facilitate the participant to attach their mouthpiece.

6.6.6. Admin will then provide instruction for how the test will run, and answer any questions the participant has.

6.6.7. If none, or all are answered, Admin will power on the machine by pressing the POWER button quickly one time (note: The power button should be hit with one quick depression/ push should be sufficient. A sustained push on the button might result in the machine to cycle back to OFF).

6.6.8. The Admin should tell participant: Please put your mouth on the machine. First you should exhale twice with your nose open and un-pinched. After that, you should pinch your nose and breathe normally until I tell you we're all set

6.6.9. Once the results are considered satisfactory, the admin should tear the piece of paper and place the result in the envelope. The admin can inform the participant to remove their mouth from the machine, but to not remove their mouthpiece.

6.7. Post-Phase A: Particle Exhalation Test Close and Clean-Up

6.7.1. Once all Exhalation Tests are complete, the Admin will ask the Participant to remove the mouthpiece from their device and place it in the bag. The admin should also ask the participant to put the cap back on the end where the mouthpiece was, and encourage them to firmly attach the cap.

6.7.2. Administrator should tear the printer receipt from the participant, and prepare it with the appropriate information:

6.7.3. The administrator should come around to the front of the protector screen, wipe down the screen, table, and chair with a sanitizing wipe.

6.7.4. Administrator will now be ready to for another participant at their station.

6.8. Phase A

6.8.1. BEFORE the participant arrives, the FEND Administrator (Admin) should prepare the station.

6.8.2. The Admin can then usher up the next participant.

6.9. Phase A: Return to Registration / Check-out 14

6.9.1. Participant returns back to the Registration Table to the "Check Out" side. Once there, the participant will hand their Nebulizer Assembly bag (with the assembly, mouthpiece, and cap inside). At the station the administrator says to the participant: "Thank you for your time. I will take your filter bag, sanitize it and store it hear for you to use again.

6.9.2. The admin will take note of the time, and on a 3x5 note card or sticker, the admin will write the time at which the participant should return (if return). The admin will write the participant's number on the card with the time, and hand it to the participant.

6.9.3. The admin will then say to the participant: Thank you so much for your time.

6.9.4. The admin will write the current time on the table with the participant's name, noting that they received the bag and that the participant completed the first cycle.

6.9.5. The admin will then spray the participant's bag with sanitizing spray (being careful to NOT spray the number written on the bag or on the sticker—so that they do not fall off). The admin places the bag in the container, in ascending number order (to facilitate the participants return.

6.10. Phase B Check-In (Phase B Repeats the Above)

6.10.1. At the end of Phase B Admin will ask for the participant's nebulizer assembly bag. The admin will sanitize the bag, by spraying the bag with alcohol cleaner, taking care to not spray the number or the sticker (which could become dislodged or wiped from the bag). If it does, the admin should rewrite the number on the bag, so that it does not lose its participant number.

6.10.2. The admin will put the bag in the bin, in ascending numeric order with the stickers and numbers in the same corner.

6.10.3. Participant may exit.

Appendix A Physical Inventory Checklist

1. Materials to Be Printed Checklist

#### About This Document

| | |
|---|---|
| **Document Name** | AEROSOLIZATION STUDY PROTOCOL - NORTH CAROLINA STUDY |
| **Document Number** | 01-0002 |
| **Version Number** | 01.06 |
| **Date Last Updated** *(See **Revision History** table on last page)* | 09/11/2020 |
| **Project Management Workstream** | Research and Development |

#### Introduction

**Context**: The Aerosol Assessment Study involves the assessment of exhaled aerosol particles among the employees of No Evil Foods, administered by the Sensory Cloud team, overseen by the PI, Dr. David Edwards. Volunteers who participate in the study will be roughly 20 minutes away from their daily work routine, to assess their exhaled aerosol particles. Volunteers will be asked to sign a consent form prior to the study that clarifies the safety and purpose of the assessment with a potential side effect of a dry mouth. Participants will be able to observe the measurement in real time.

**Goals and Outcomes:** The goal of the study is to determine exhaled aerosol variation between subjects

### Aerosol Assessment Study Protocol

#### 2. Scope

##### 2.1. This process document contains information on the following:

2.1.1.Suggested protocol for the measurement of exhaled and emitted aerosolized particles, by size of particles

2.1.2.This version of the study focuses on the *variations of particle aerosolization in participants over the course of multiple, closely timed measurements* (e.g., daily over the course of a week, with multiple measurements taken each day).

2.1.2.1.1.I.e., there are two assessments performed

2.1.2.2.(2) there is one assessment performed

2.1.2.3.Additionally, this study design varies is other ways:

2.1.2.4.This study is meant to be *administered for a single household at a given time*, given the risk of communicable illness, like COVID-19. Because of this, the study should be done for all individuals willing to participate from that household, and all participants should be tested for COVID-19. The study can be repeated, but *separately* with cleaned equipment for *other participating households* at *different date*

2.1.2.5.The participants will also NOT participate in a control administration with saline.

2.1.2.6.Test administrators will track the participant's COVID-19 status (i.e., COVID-positive or COVID-negative), noting the days from the onset of COVID-like symptoms (i.e., Day 0

2.1.2.6.1.The timing of same-day measurement administrations will be measured

2.1.3.Suggested directions for participants and administrators to effectively and consistently administer this process and inform participants of the test's mechanics to aid their understanding and successful participation of study requirements

2.1.4.Required inventory list of equipment, supporting inventory, and study administration materials for successful deployment (see Appendix A)

#### 3. References

The following documents are referred to in the text in such a way that some or all of their content constitutes requirements of this document. For dated references, only the edition cited applies. For undated references, the latest edition of the referenced document (including any amendments) applies.

3.1. **Trial Flow Diagram:** Trial Flow Layout Setup

#### 4. Terms and definitions

**For the purposes of this document, the following terms and definitions apply.**

##### 4.1. Nebulizer Kits

This is the apparatus that allows the exhaled particles from the participant's airway to be counted by the particle counter. One complete kit is composed of five pieces: T-Connector valve; Flexible connector Tube; HEPA Filter Assembly; Participant's Mouthpiece, T-Connector Valve Cap (typically red)

##### 4.2. HEPA Filters and HEPA Filter Assembly

This device filters the ambient air of particles, so that the particle counter is measuring only those particles exhaled from the participant. Because it serves this purpose, the HEPA filter and all joints and connections for between the parts of the Nebulizer kit must be tight and secure. This is also why the Particle Counter Administrator must perform a baseline to ensure that the seals are tight and the HEPA filter is filtering properly. See study administration protocol, Particle Counter, to see the minimum required readings for the HEPA Filter Baseline Test.

##### 4.3. T-Connector Valve Caps (Used for HEPA Filter Testing / Baseline)

Every Nebulizer Kit includes a cap that is a small convex plastic cap, that fits snugly over the ends of the T-Connector Valve. When properly connected, one end of the top of the T-Connector Valve is connected to the particle counter; the bottom valve is connected to the Joint Connector Tube (which connects to the HEPA filter). The cap will be fitted to the other top valve of the T-Connector Valve before the participant attaches their mouthpiece to perform the baseline test. The cap seals tightly to the T-Connector Valve blocking airflow through that side.

##### 4.4. Particle Counter Receipt Paper

Each particle counter machine measures counts over a 6-second period. The results of this count are measured in real time, and the results are captured on physical thermal print paper (much like a cash register at a store). Each participant will have ONE printout of their results over the ~2-3 minutes of breathing (with results measured for each 6 second interval), for each administration of the particle test. These printed results are the only physical record of the participant's results, and therefore are critical to capture, organize, and store securely. The machines should have rolls of paper in the device, with many extras nearby.

#### 5. Roles and responsibilities

For the purposes of this document, the following roles apply with the responsibilities listed besides each.

##### 5.1. Study Project Manager

Managing all study related logistics, collaborating cross-functionally within Sensory Cloud and externally with the partner organization to ensure all information/materials required for the study are arranged for in time.

##### 5.2. Study Site Point of Contact (POC)

Responsible to manage relationships with the site owners, hosts, or "client" (depending on the nature of the test administration), and to oversee and engage other SC team members to drive successful execution

##### 5.3. Registration Administrator

Responsible for staffing the Registration Station, which includes the following: Checking in participants, assigning participants a Participant Number, confirming participants have signed Informed Consent Waiver, provision participants with their Ziplock bag, mouthpiece, and cap; direct them to the particle test station, receive them when they complete the aerosolization station; collect, sanitize, and store (in numeric order) participants nebulizer kit bags; write down and inform participant of their next test; and finally collect their nebulizer kits (once again sanitizing) and thanking participant for their time.

##### 5.4. Particle Test Administrator

Responsible for staffing the Particle Counter Machines, facilitating the participant through the particle test. BEFORE each participant is received at the counter, the Administrator should wipe/spray down the chair, separator screen, and table; secure an UNUSED nebulizer assembly (with cap) to the particle counter; and check the machine has sufficient paper. WHEN PARTICIPANT ARRIVES, Administrator will welcome participant; instruct the participant about the process they will take; guide the participant through the exhalation test process (and correct for any errors); mark the receipt paper with any errors; thank the participant for their time; direct them to the Aerosolization station (or the Registration Table). AFTER Participant leaves, the Administrator will clean the station, and repeat

##### 5.5. Runner to Coordinate with Participants

Responsible for informing participants to come to the registration desk and make sure that only the set number of participants (i.e., 3) are brought into the testing space for registration at a single time. This is part of the protocol to reduce the risk of inter-personal transmission.

##### 5.6. Trial participants

Willful and informed participant in the study, and for this study are parts of the same household, given the presence and tracking of COVID-19 positivity within the household. Participants must sign consent form, provide basic data as solicited on the Informed Consent Form, and follow the instruction of all study administrators.

### 6. Process

#### 6.1. Pre-Work / Things Done in Advance

6.1.1.Gather/Acquire all materials required as per materials list (see full inventory list in Appendix A)

6.1.1.1.Gather materials in inventory

6.1.1.2.List out materials in transit or yet to be purchased

6.1.1.3.Place orders/ track delivery

6.1.1.4.Ensure all travel arrangements made

6.1.2.Organize Dry-Run Before Trial

6.1.2.1.Setup room exactly as per layout

6.1.2.2.Have all materials laid out next to each station as planned

6.1.2.3.Run-through the entire process with the team

#### 6.2. Visit Study Location

6.2.1.Room Setup for Trial

6.2.1.1.Set up each trail station as described below. There are 4 stations for each administration, all trial stations with materials (***Note***: This list is illustrative only; please refer to Appendix A for complete inventory pack list)

6.2.1.1.1.Registration/Check-in/Check-out Station

6.2.1.1.1.1.Consent Forms

6.2.1.1.1.2.Pens

6.2.1.1.1.3.Labels for allotting number to participants

6.2.1.1.1.4.Mouthpiece in resealable bags

6.2.1.1.1.5.Participant check-in/check-out list

6.2.1.1.1.6.Bin to store consent forms

6.2.1.1.1.7.Bin to store sealed kit bags

6.2.1.1.2.Particle Testing Station

6.2.1.1.2.1.Particle Counter Machine with plexiglass separator screen

6.2.1.1.2.2.Pre-assembled filter kit

6.2.1.1.2.3.Envelopes to store receipts (i.e., particle test results)

6.2.1.1.2.4.Fine point pens

6.2.1.1.2.5.Bin to store numbered envelopes in

6.2.1.1.2.6.Extra receipt roles for particle counter

6.2.1.1.2.7.Dry- Run at study location

6.2.1.1.2.8.Take them through all stations and explain the testing cycle

6.2.1.2.Locate materials and resources to be organized and offered by study host location

6.2.1.2.1.Runner to help communicate with employees

6.2.1.2.2.Identify go-to person for any logistical questions

#### 6.3. Overview of Particle Exhalation Baseline

6.3.1.Participants will proceed through several measurements, taking place over several days. Please defer to the study Principle Investigator (i.e., PI) to determine the final protocol for the specific study administration.

6.3.2.In this study, participants within the same household will be tested at the same sequence.

6.3.2.1.)

#### 6.4. Phase A: Registration

*6.4.1.*Participant comes into the test administration room. The Registration Administrator (Admin) welcomes them: *Thank you for making time to participate in this study. Can I please have your First and Last Names?*

6.4.1.1.Admin completes the Online Form (or if no internet connection is available, enter into pre-formatted table) with the participant's name and the number they are assigned. Each participant should be assigned a number; the numbers do not have to be assigned in ascending order. However, each participant's number must be documented with their name in order to track the completion of Informed Consent Waivers and the completion of all 4 tests of the study protocol.

6.4.1.2.The Admin will put one sticker with the participant's number on their bag, and another on an envelope. The third sticker will be put on the participant's shirt

6.4.2.Admin should also ask the participant if they have **tested positive for COVID-19**. If so, the administrator should make a note, and ask when the participant was tested, and when their symptoms were first observed.

6.4.2.1.Admin should write these facts legibly on the consent form, rather than leaving it for the participant to complete themselves.

*6.4.3.*Admin says *Have you had a chance to review and sign our informed consent waiver?*

*6.4.3.1.*(If not) *Here is a consent form that you can go through and sign on page 2. On the last page, you will be required to fill in some basic information about yourself such as age, gender, smoking habits and history of lung disease.*

6.4.3.2.Participant should sign and complete their consent form. If there is another participant behind them waiting, kindly ask the participant to step aside while the next participant checks in.

6.4.3.3.(If yes) *Thank you for completing the form*

*6.4.4.*Overview of process: *The test process will take about 20 minutes. You will be asked to breathe into the particle counter machine using your own new filter kit assembly. We want to measure the number and size of particles our participants give off, when you breath or create when talking.*

*6.4.5.*Participant is handed their Ziploc bag with the mouthpiece inside. Admin says: *At the next station, our Administrations will guide you through how to use the particle counter. To reduce the risk of any transmission of illness, only you will touch the component that will go in your mouth. This bag was packaged by our team, on clean surfaces, wearing gloves, to make sure it is clean for you.*

*6.4.5.1.At the station, the administrator will have attached your filter to the machine. They will guide you through how to attach the mouthpiece you see in this bag.*

*6.4.6. The particle count test will run for about 5 minutes. When you are done, our team will direct you back to me at the registration desk. At that time, I will collect and sanitize the bag with your Nebulizer Assembly (including your mouthpiece) to be kept by the desk. I will also let you know what time to be back at the registration desk.*

*6.4.7. Do you have any questions before we begin?*

6.4.8. Admin will hand the participant three items

6.4.8.1. Bag with the sticker (with participant's number on it), and a sharpie-written number on the bag in the corner

6.4.8.2. Envelope with the participant's number and name in the corner

6.4.8.3. Sticker with the participant's number and name to put on their shirt / uniform

*6.4.9.* Admin will say: *Please proceed to the next station (indicating where it is). When you arrive, please give your envelope to the administrator at the station. It will have been sanitized before you arrive.*

6.4.10. Admin can then proceed to address the following participant in line.

#### 6.5. Phase A: Particle Exhalation

6.5.1. Before the participant arrives, the admin will come around from the particle counter and the separator screen. They will sanitize with wipes the seat, screen, and tabletop.

6.5.1.1. Station administrator (admin) will attach a pre-assembled nebulizer filter kit (with the red cap in place; stored in a bin at the station)

6.5.1.2. They will also have to remove the two labels that cover the HEPA filter (i.e., blue hard plastic box) and discard the sticker labels. The sticker is most easily removed by lifting a corner of the sticker with a fingernail and peeling slowly back away from the hard plastic container

*6.5.2.* When the participant arrives, the admin will direct them to kindly sit down in the chair in front of the machine. Admin: *Thank you for participating! At this station, we will measure the number and size of the particles you produce while breathing. At the registration table, you were given a bag and an envelope. Can I please have the envelope?*

*6.5.2.1.* Participant should then give their envelope to the administrator. Admin: *Thank you*

*6.5.3. To begin, I will activate the machine so you get a sense for how it sounds and work. Right now, you will see the red cap is sealing where your mouthpiece and mouth will go. This will help us make sure the machine is properly working—and give us a baseline against which to measure your particle counts and sizes.*

6.5.3.1. Admin will activate the machine by quickly pressing the POWER button, and hitting start (the yellow icon in the bottom LEFT HAND corner of the screen). This activates the vacuum motor and will begin to draw air in through the HEPA filter. Admin will run the machine so at least 4 printed intervals (i.e., a 6-second interval printed on a receipt).

6.5.4. During this test, if the >0.3 µm count (the lowest of the four numbers) on the particle counter machine screen is > 40 for more than ten seconds, AND/OR if the>0.3 µm count is variable (i.e., the count jumps), let the machine run for 20 seconds (~3-4 6-second intervals). If the count continues to exceed the number above, or is variable, the admin should take the following steps to remediate:

6.5.4.1. Slowly and with small movements, move around with nebulizer assembly and the particle machine connector tube to see if the numbers jump more. If the numbers do jump while the assembly is moved, it is likely that there is a leak or an insecure connection between some components

6.5.4.2. First come around the plexiglass screen and make sure that every seal between Nebulizer Assembly parts is tight. This is especially true for the red cap, which may have become loosened during transit.

6.5.4.3. Move around the connections between the components of the assembly, to migrate them a little bit relative to one another. If the numbers on the counter jump, there are joints that need to be tightened.

6.5.4.4. Check the connection between the nebulizer kit and the machine connection nozzle. Since this is attached by hand, it can require some force to make a seal.

6.5.4.5. Restart the machine and check particle counts; if the >0.3 µm count on the particle counter machine screen is > 40 do the following:

6.5.4.6. Disconnect the entire Nebulizer Assembly and discard. Retrieve a new one from the bin, and redo the test. If numbers still exceed threshold, repeat the whole baseline test.

6.5.5. If the numbers **do not exceed** thresholds, the Admin should turn off the counter (but hitting STOP on the bottom left hand yellow button) for machine. The test can begin. The Admin will then facilitate the participant to attach their mouthpiece.

6.5.5.1. Admin: *For safety, please sanitize your hands before we begin. Please remove the red cap from the Nebulizer Assembly and place if back in your bag.*

*6.5.5.2. Now please remove your mouthpiece from the bag and attach it firmly around the valve opening where the red cap was. Kindly press firmly; it should be a snug fit—and might need more force than you think.*

6.5.5.3. Participant follows instructions; Admin will visually inspect the mouthpiece from behind the screen. If the participant needs any assistance, the Admin will come around to help them, wearing mask, face shield, and gloves.

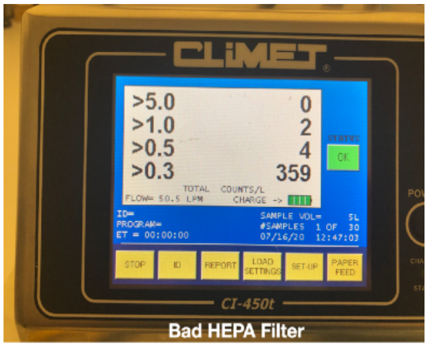

Bad HEPA Filter

6.5.6. Admin will then provide instruction for how the test will run, and answer any questions the participant has.

6.5.6.1. Admin: *As you might remember, this test measures the number and size of particles you breath out. While it might feel weird, we actually want you to breath as normally as possible, breathing in and out of the mouthpiece.*

*6.5.6.2. The machine measures the particle size and count in 6 second intervals, so we will ask you to keep your mouth on the mouthpiece, breathing normally for up to 3 minutes. A few things to keep in mind:*

*6.5.6.3. Keep a tight but comfortable seal between your lips and the mouthpiece until I tell you the test is complete. If the seal is broken, we may have to restart the test.*

*6.5.6.4. We will require that you pinch your nose for that whole time as well, so that all air inhaled comes through the HEPA filter. This is to make sure particles above a certain size to not enter the machine—and that all your exhaled air goes into the machine (and not out of your nose). If you have any issues with this, please let me know.*

*6.5.6.5. When you first put your mouth over the mouthpiece, take two deep inhales and exhalations with your nose UNPINCHED. These breaths will clear your lungs of air that is in your lungs from before the filtered air enters. These should be deep exhalations.*

*6.5.6.6. Once those two breaths are complete, please proceed to pinch your nose, and breath normally while maintaining the pinch on your nose.*

*6.5.6.7. Again, we will ask you to breath normally for the whole 2-3 minutes. Please do not stop or remove your mouth or stop pinching your nose until I let you know. Do you have any questions?*

6.5.6.8. Admin will pause to answer any questions the participant has.

6.5.7. If none, or all are answered, Admin will power on the machine by pressing the POWER button quickly one time (note: The power button should be hit with one quick depression/push should be sufficient. A sustained push on the button might result in the machine to cycle back to OFF).

6.5.8. The Admin should tell participant: *Please put your mouth on the machine. First you should exhale twice with your nose open and un-pinched. After that, you should pinch your nose and breathe normally until I tell you we're all set.*

6.5.8.1. The participant should put their lips on the mouthpiece, and take two deep exhalations. The Admin should then make sure the participant pinches their nose, and continues to breathe.

6.5.8.2. During the 2-3 minutes, the Admin should periodically encourage the participant and remind them that they are doing well. The admin should **NOT** ask any questions or make jokes that might encourage the participant to remove their mouth to respond or react in any way. Consider the following:

*6.5.8.2.1. You are doing great; exactly as you're doing*
*6.5.8.2.2. Thanks for keeping your nose pinched; this is going perfectly.*
*6.5.8.2.3. Nice work; just keep breathing normally. This is exactly how it should work.*

6.5.8.3. After 60 seconds, the Admin should check the printing receipt and confirm that the results are satisfactory. If they are, the machine motor should be deactivated. Administrators should confirm that the receipt paper indicates the following data were acquired:

6.5.8.3.1. 5-8x 6-second interval printed results with relatively consistent particle counts across all four size categories

6.5.8.3.2. If any intervals appear to "jump" with high particle counts, the admin should continue the test for another 5-6x 6-second interval results. The "jump" can often be attributed to latent atmospheric (i.e., unfiltered) air within the lungs being expelled.

6.5.8.4. At no point should the participant be allowed to remove their lips from the machine, or to un-pinch their nose after the two initial breaths.

6.5.9. Once the results are considered satisfactory, the admin should tear the piece of paper and place the result in the envelope. The admin can inform the participant to remove their mouth from the machine, but to not remove their mouthpiece.

#### 6.6. Post-Phase A: Particle Exhalation Test Close and Clean-Up

6.6.1. Once all Exhalation Tests are complete, the Admin will ask the Participant to remove the mouthpiece from their device and place it in the bag. **The admin should also ask the participant to put the cap back on the end where the mouthpiece was, and encourage them to firmly attach the cap.**

6.6.1.1. Administrator should then come around the separator to remove the Nebulizer Assembly from the nozzle, and hand it to the participant.

6.6.1.2. Participant is asked to put the remaining filter assembly handed over by the administrator in the resealable bag. The admin will thank the participant for their time and direct them to the following station (FEND administration)

6.6.2. Administrator should tear the printer receipt from the participant, and prepare it with the appropriate information:

6.6.2.1. On the paper, the admin should write the participant's number (from the envelope).

6.6.2.2. Administrator draws line on receipt paper printed from particle counter where the particle exhalation stabilizes (e.g., after the two initial exhalations when the particle counts have ceased to "jump"). The printed particle counts above this line will be considered baseline, while those printed number below will be considered the test administration.

6.6.2.3. The admin should also write on the receipt paper the name of the study phase for which the receipt:

6.6.2.3.1. Baseline

6.6.2.4. The receipt paper should be folded (NOT ROLLED) and slipped into the envelope, in order to keep the receipts as flat as possible. Do NOT SEAL the envelope.

6.6.2.5. Administrator will put the participant's envelope in the box for their station (NOTE: each machine station should have its OWN box of envelopes). The envelopes should be kept in number order, facing the same way, to facilitate finding the envelopes later

6.6.3. The administrator should come around to the front of the protector screen, wipe down the screen, table, and chair with a sanitizing wipe.

6.6.3.1. The admin should also attach another (as of yet unused) Nebulizer Assembly from the bin (with the cap attached) to the machine nozzle. The administrator should check and tighten the connections between joints in the Nebulizer Assembly and the machine nozzle, and in the joints in the Nebulizer Assembly (e.g., on the T-Connector Valve; the HEPA filter; the joint connector tube.

6.6.4. Administrator will now be ready to for another participant at their station.

#### 6.7. Phase A: Return to Registration / Check-out

*6.7.1.* Participant returns back to the Registration Table to the "Check Out" side. Once there, the participant will hand their Nebulizer Assembly bag (with the assembly, mouthpiece, and cap inside). At the station the administrator says to the participant: "*Thank you for your time. I will take your filter bag, sanitize it and store it hear for you to use it again.*

6.7.2. The admin will take note of the time, and on a 3x5 note card or sticker, the admin will write the time at which the participant should return (i.e., the current time + 30 mins). The admin will write the participant's number on the card with the time, and hand it to the participant.

*6.7.3.* The admin will then say to the participant: *Thank you so much for your time. We kindly ask that you please return for the next test (indicating to the card with the time and number). .*

*6.7.3.1. When you return to the test room at the designated time, please come straight to the registration desk, and hand this card (indicating to the card with the time and participant number) back to me or the person at this station. We will sanitize and keep this bag safely at our station.*

*6.7.3.2. Do you have any questions?*

6.7.3.3. If the participant has any questions, the admin will answer them to the best of their ability

6.7.4. The admin will write the current time on the table with the participant's name, noting that they received the bag and that the participant completed the first cycle.

6.7.5. The admin will then spray the participant's bag with sanitizing spray (being careful to NOT spray the number written on the bag or on the sticker—so that they do not fall off). The admin places the bag in the container, in ascending number order (to facilitate the participants return.

#### 6.8. Phase B Check-In (Phase B Layout Above)

*6.8.1.*Participant comes into the test administration room for the next test.

6.8.1.1.Admin completes the Google Form or table (if no internet is available) the table with the participant's name and the number they are assigned. This will allow the Admin to track the participant's start time for the Phase B Assessment.

6.8.1.2.If the participant does not have their sticker on their clothes with their participant number, OR the time card with the participant number on it, the Admin should look at the Google Form output to cross-reference the participant's name with their participant number.

6.8.1.3.Once the admin finds the number, the admin should retrieve the participant's nebulizer assembly bag from the bin (they should be in numeric order).

6.8.1.4.Administrator checks that participant is wearing their number tag on their shirt before they enter the testing room

*6.8.1.5.The admin says to the participant: This next part of the study will take less time than the previous administration. You will proceed to the breathing station directly once again.*

*6.8.1.6. The breathing test will be exactly the same as you performed before the wait, except that this time you will hand the bag to the administrator of that station, and he will attach the nebulizer to the machine. You will once again attach your own mouthpiece.*

*6.8.1.7.Before you begin, do you have any questions?*

6.8.2.Admin will answer any questions. If there are no questions or all are answered, the admin will direct the participant to the breathing station.

*6.8.2.1.*Admin will say: *Please go to the same machine and administrator that facilitated you through the first test.*

#### 6.9. Phase B: Particle Exhalation Test 2

6.9.1.Before the participant arrives, the admin will come around from the particle counter and the separator screen. They will sanitize with wipes the seat, screen, and tabletop. The administrator should put on clean gloves, discarding any that they were wearing before.

*6.9.2.*When the participant arrives, the admin will direct them to kindly sit down in the chair in front of the machine. Admin: *Thank you for participating! At this station, we will again measure the number and size of the particles you produce while breathing. At the registration table, you were given a bag. Can I please have the bag with your nebulizer assembly?*

*6.9.2.1.*Participant should then give their bag to the administrator. Admin: *Thank you*

6.9.2.2.The admin should reference the number on the bag, and retrieve the corresponding envelope from the box of envelopes and receipts. The admin should place the envelope on the table.

6.9.2.3.The admin will remove the nebulizer assembly with the red cap on from bag, leaving the mouthpiece in the bag. The admin should double check that the two sticker labels are removed from the hard plastic exterior of the HEPA filter; these should have been removed during the first exhalation test. The admin should secure the nebulizer kit to the particle counter nozzle. The admin should make sure that the joints are tight, and that the red cap is secure.

*6.9.3.*Admin will say: *Same as before, I will activate the machine so you get a sense for how it sounds and works. Right now, you will see the red cap is sealing where your mouthpiece and mouth will go. This will help us make sure the machine is properly working—and give us a baseline against which to measure your particle counts and sizes.*

6.9.3.1.Admin will activate the machine by quickly pressing the POWER button, and hitting start (the yellow icon in the bottom LEFT HAND corner of the screen). This activates the vacuum motor and will begin to draw air in through the HEPA filter. Admin will run the machine so at least 4 printed intervals (i.e., a 6-second interval printed on a receipt).

6.9.3.2.During this test, if the >0.3 µm count (the lowest of the four numbers) on the particle counter machine screen is > 40 for more than ten seconds, AND/OR if the>0.3 µm count is variable (i.e., the count jumps), let the machine run for 20 seconds (~3-4 6-second intervals). If the count continues to exceed the number above, or is variable, the admin should take the following steps to remediate:

6.9.3.3.Play with the tube to see if the numbers jump more; making it clear that there is a leak

6.9.3.4.First come around the plexiglass screen and make sure that every seal between Nebulizer Assembly parts is tight. This is especially true for the red cap, which may have become loosened during transit.

6.9.3.5.Move around the connections between the components of the assembly, to migrate them a little bit relative to one another. If the numbers on the counter jump, there are joints that need to be tightened.

6.9.3.6.Check the connection between the nebulizer kit and the machine connection nozzle. Since this is attached by hand, it can require some force to make a seal.

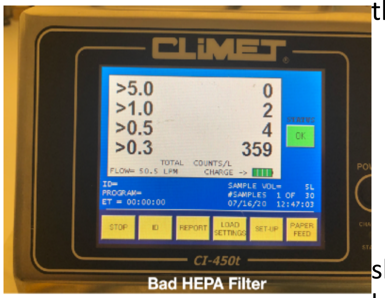

6.9.3.7.Restart the machine and check particle counts; if the >0.3 µm count on the particle counter machine screen is > 40 do the following:

6.9.3.8.Disconnect the entire Nebulizer Assembly and discard. Retrieve a new one from the bin, and redo the test. If numbers still exceed threshold, repeat the whole baseline test.

6.9.4.If the numbers **do not exceed** thresholds, the Admin should turn off the counter (but hitting STOP on the bottom left hand yellow button) for machine. The test can begin. The Admin will then facilitate the participant to attach their mouthpiece.

*6.9.4.1.*Admin: *For safety, please sanitize your hands before we begin. Please remove the red cap from the Nebulizer Assembly and place if back in your bag.*

*6.9.4.2.Now please remove your mouthpiece from the bag and attach it firmly around the valve opening where the red cap was. Kindly press firmly; it should be a snug fit—and might need more force than you think.*

6.9.4.3.Participant follows instructions; Admin will visually inspect the mouthpiece from behind the screen. If the participant needs any assistance, the Admin will come around to help them, wearing mask, face shield, and gloves.

6.9.5.Admin will then provide instruction for how the test will run, and answer any questions the participant has.

*6.9.5.1.*Admin: *As you might remember, this test measures the number and size of particles you breath out. While it might feel weird, we actually want you to breath as normally as possible, breathing in and out of the mouthpiece.*

*6.9.5.2.The machine measures the particle size and count in 6 second intervals, so we will ask you to keep your mouth on the mouthpiece, breathing normally for up to 3 minutes. A few things to keep in mind:*

*6.9.5.3.Keep a tight but comfortable seal between your lips and the mouthpiece until I tell you the test is complete. If the seal is broken, we may have to restart the test.*

*6.9.5.4.We will require that you pinch your nose for that whole time as well, so that all air inhaled comes through the HEPA filter. This is to make sure particles above a certain size to not enter the machine—and that all your exhaled air goes into the machine (and not out of your nose). If you have any issues with this, please let me know.*

*6.9.5.5.When you first put your mouth over the mouthpiece, take two deep inhales and exhalations with your nose UNPINCHED. These breaths will clear your lungs of air that is in your lungs from before the filtered air enters. These should be deep exhalations.*

*6.9.5.6.Once those two breaths are complete, please proceed to pinch your nose, and breath normally while maintaining the pinch on your nose.*

*6.9.5.7.Again, we will ask you to breath normally for the whole 2-3 minutes. Please do not stop or remove your mouth or stop pinching your nose until I let you know. Do you have any questions?*

6.9.5.8.Admin will pause to answer any questions the participant has.

6.9.6.If none, or all are answered, Admin will power on the machine by pressing the POWER button quickly one time (note: The power button should be hit with one quick depression/push should be sufficient. A sustained push on the button might result in the machine to cycle back to OFF).

6.9.7.The Admin should tell participant: *Please put your mouth on the machine. First you should exhale twice with your nose open and un-pinched. After that, you should pinch your nose and breathe normally until I tell you we're all set.*

6.9.7.1.The participant should put their lips on the mouthpiece, and take two deep exhalations. The Admin should then make sure the participant pinches their nose, and continues to breathe.

6.9.7.2.During the 2-3 minutes, the Admin should periodically encourage the participant and remind them that they are doing well. The admin should **NOT** ask any questions or make jokes that might encourage the participant to remove their mouth to respond or react in any way. Consider the following:

*6.9.7.2.1.You are doing great; exactly as you're doing*

*6.9.7.2.2.Thanks for keeping your nose pinched; this is going perfectly.*

*6.9.7.2.3.Nice work; just keep breathing normally. This is exactly how it should work.*

6.9.7.3.After 60 seconds, the Admin should check the printing receipt and confirm that the results are satisfactory. If they are, the machine motor should be deactivated. Administrators should confirm that the receipt paper is the following:

6.9.7.3.1.5-8 6-second interval results with relatively consistent particle counts

6.9.7.3.2. If any intervals appear to “jump” with high particle counts, the admin should continue the test for another 5-6 6-second interval results. The “jump” can often be attributed to latent atmospheric (i.e., unfiltered) air within the lungs being expelled.

6.9.7.3.3. At no point should the participant be allowed to remove their lips from the machine, or to un-pinch their nose after the two initial breaths.

6.9.7.4. Once the results are considered satisfactory, the admin asks the Participant to remove the mouthpiece from their device and place it in the bag. **The admin should also ask the participant to put the cap back on the end where the mouthpiece was, and encourage them to firmly attach the cap.**

6.9.7.5. Administrator should then come around the separator to remove the Nebulizer Assembly from the nozzle, and hand it to the participant.

6.9.7.6. Participant is asked to put the remaining filter assembly handed over by the administrator in the resealable bag. The admin will thank the participant for their time and direct them to the following station (FEND administration)

6.9.8. Administrator should tear the printer receipt from the participant, and prepare it with the appropriate information:

6.9.8.1. On the paper, the admin should write the participant’s number (from the envelope).

6.9.8.2. Administrator draws line on receipt paper printed from particle counter where the particle exhalation stabilizes (e.g., after the two initial exhalations when the particle counts have ceased to “jump”). The printed particle counts above this line will be considered baseline, while those printed number below will be considered the test administration.

6.9.8.3. The admin should also write on the receipt paper the name of the study phase for which the receipt

6.9.8.4. The receipt paper should be folded (NOT ROLLED) and slipped into the envelope, in order to keep the receipts as flat as possible. Do NOT SEAL the envelope.

6.9.8.5. Administrator will put the participant’s envelope in the box for their station (NOTE: each machine station should have its OWN box of envelopes). The envelopes should be kept in number order, facing the same way, to facilitate finding the envelopes later

6.9.9. The administrator should come around to the front of the protector screen, wipe down the screen, table, and chair with a sanitizing wipe.

6.9.9.1. The admin should also attach another (as of yet unused) Nebulizer Assembly from the bin (with the cap attached) to the machine nozzle. The administrator should check and tighten the connections between joints in the Nebulizer Assembly and the machine nozzle, and in the joints in the Nebulizer Assembly (e.g., on the T-Connector Valve; the HEPA filter; the joint connector tube.)

6.9.10. Administrator will now be ready to for another participant at their station.

#### 6.10. Close Out

6.10.1. Admin: Thank you for participating in the study.

6.10.1.1. (If Day 1) We appreciate that you can come back in tomorrow at this same location and approximate time, to have the control study completed.

6.10.2. Admin will ask for the participant's nebulizer assembly bag. The admin will sanitize the bag, by spraying the bag with alcohol cleaner, taking care to not spray the number or the sticker (which could become dislodged or wiped from the bag). If it does, the admin should rewrite the number on the bag, so that it does not lose its participant number.

6.10.3. The admin will put the bag in the bin, in ascending numeric order with the stickers and numbers in the same corner.

6.10.4. Participant may exit.

### Appendix A

### Physical Inventory Checklist

| Item | Quantity | Packed? | Box # |
|---|---|---|---|
| **Particle Counter** | 3x | | |
| **Receipt Paper Rolls** | 3x/ machine | | |
| **Nimbus units** | 3x/ machine | | |
| **6x AAA Batteries / Machine** | 18x | | |
| **HEPA Filter Assembly** | 150x | | |
| **T-Connector Valve** | " | | |
| **Joint Tube Connector** | " | | |
| **Mouthpieces** | 150x | | |
| **T-Connector Valve Cap (Red)** | 150x | | |
| **HEPA Filters** | 150 | | |
| **Plexiglass Separator Screen** | 3x | | |
| **Ziplock Bags (2 Gallon size)** | 150 | | |
| **Staples Laser/Inkjet Address Labels, 1" x 2 5/8", White, 30 Labels/ Sheet, 100 Sheets/Box (18057/SIWO100)** *Printed with ascending numbers at left side of stickers (to the max number of anticipated participants +20%), no names* | 3x/ participants | | |
| **Staples Thermal Paper Rolls (2.25"x50') –** (816613) | 10x/ machine | | |
| **Sharpie Felt Pens, Fine Point, Black Ink** | 3x / Machine + 4 for registration | | |
| **Sharpie Original (fat tip) pen, Black Ink** | 4x | | |
| **Ballpoint Pens** | 10 | | |
| **Large Volume Alcohol Hand Sanitizer with Hand Pump** | 4x/ machine | | |
| **Stapler** | 2 | | |
| **Ballpoint Pens** | 10 | | |
| **KN-95 Masks** | 8x/ administrator | | |
| **Face Shields** | 1x administrator | | |
| **Latex gloves (Large)** | 2 boxes | | |
| **Envelopes with seal (to fit 8.5x11" paper)** | 3x/ participants | | |
| **8.1 Liter Snap Lid Storage Bin, Clear (8.1L CL) for Envelops** | 1x | | |
| **64L Hard Plastic (clear) container with snap-lock solid lid** | 5x | | |
| **Kleenex** | 2 boxes | | |

#### 1. Materials to Be Printed Checklist

| Item | Quantity | Packed? | Box # |
|---|---|---|---|
| **Staples Laser/Inkjet Address Labels, 1" x 2 5/8", White, 30 Labels/ Sheet, 100 Sheets/Box (18057/SIWO100)**<br>*Printed with ascending numbers at left side of stickers (to the max number of anticipated participants +20%), no names* | 3x/ participants | | |
| **Informed Consent Waiver + Participant Data Sheet**<br>*Should print for # of participants + 20%* | 150x | | |
| **Detailed Study Administration Schedule** | 12x | | |
